# Managing bed capacity and timing of interventions: a COVID-19 model considering behavior and underreporting

**DOI:** 10.1101/2022.03.29.22273148

**Authors:** Victoria May P. Mendoza, Renier Mendoza, Youngsuk Ko, Jongmin Lee, Eunok Jung

## Abstract

**Introduction:** At the start of the pandemic, the Philippine capital Metro Manila was placed under a strict lockdown termed Enhanced Community Quarantine (ECQ). When ECQ was eased to General Community Quarantine (GCQ) after three months, healthcare systems were soon faced with a surge of COVID-19 cases, putting most facilities at high or critical risk and prompting a return to a stricter policy.

**Methods:** We developed a mathematical model considering behavior changes and underreporting to represent the first major epidemic wave in Metro Manila. Key parameters were fitted to the cumulative cases in the capital from March to September 2020. A bi-objective optimization problem was formulated that allows easing of restrictions at an earlier time and minimizes the necessary additional beds to ensure sufficient capacity in healthcare facilities once ECQ was lifted.

**Results:** If behavior was changed one to four weeks earlier before GCQ, then the cumulative number of cases can be reduced by up to 55% and the peak delayed by up to four weeks. Increasing the reporting ratio during ECQ threefold may increase the reported cases by 23% but can reduce the total cases, including the unreported, by 61% on June 2020. If GCQ began on May 28, 2020, 48 beds should have been added per day to keep the capacity only at high-risk (75% occupancy). Among the optimal solutions, the peak of cases is lowest if ECQ was lifted on May 20, 2020 and with at least 56 additional beds per day.

**Conclusion:** Since infectious diseases are likely to reemerge, the formulated model can be used as a decision support tool to improve existing policies and plan effective strategies that can minimize the socioeconomic impact of strict lockdown measures and ensure adequate healthcare capacity.

## 1. Introduction

Shortly after the first local transmissions of the coronavirus disease 2019 (COVID-19) in the Philippines were confirmed, the government imposed social distancing policies, termed community quarantines, which were largely implemented by the police and military [1, 2]. By March 30, 2020, the country only had six laboratories that accommodated up to 1000 tests daily [3]. Contact tracing began slowly due to insufficient number of contact tracers [4]. Testing capacity was increased to about 35000 tests daily by the end of September 2020 [5]. From April to September 2020, the weekly positivity rate ranged between 4.5 and 28%, higher than the 5% threshold set by the World Health Organization during this time [6, 7].

The Philippines’ capital region, Metro Manila, comprised around 12% of the country’s population in 2020 [8]. Metro Manila was placed under *Enhanced Community Quarantine* (ECQ) on March 16, 2020 [9]. Under ECQ, movement was restricted to essential goods and services. Public transportation and mass gatherings were suspended [2]. People were encouraged to work from home and businesses were advised to do transactions online [1]. After two months, ECQ was replaced by the *Modified Enhanced Community Quarantine* (MECQ), a transition phase before easing further to *General Community Quarantine* (GCQ). On June 1, 2020, Metro Manila was placed under GCQ, where public transportation and other establishments, except those for leisure, were allowed to operate [2]. A surge in the number of cases occurred from July to August 2020 and consequently, Metro Manila was again placed under MECQ. During this time, the utilization of ICU, isolation, and ward beds in Metro Manila reached 77%, 74%, and 84%, respectively, placing most facilities on critical or high-risk and prompting 80 medical societies, representing 80000 doctors and a million nurses to demand a ‘timeout’ [10, 11]. On August 19, 2020, Metro Manila returned to GCQ [12]. By the end of September 2020, 53% of the 309303 total confirmed cases in the Philippines belonged to Metro Manila [5].

Because of the lack of vaccines and limited antiviral therapies during the early phase of the COVID-19 pandemic, NPIs such as wearing of masks, school and workplace closures, and travel restrictions were crucial disease control measures. In the Philippines, compliance to policies was not only prompted by public heath campaigns, but also driven by uncertainty and anxiety about the disease, and fear of getting reprimanded by the authorities [13, 14, 15]. Some of those who got infected suffered stigma and were blamed for not following the protocols [16, 17]. A study among low income households in the Philippines done in the early phase of the pandemic reported that 66% of respondents who might experience symptoms considered staying at home instead of seeking medical attention [13].

Non-pharmaceutical interventions and behavior change have been incorporated into mathematical models of COVID-19 [18, 19, 20, 21]. In this study, we extend the SEIQR model developed by Kim et al. [21] that includes a compartment for behavior-changed susceptible individuals. We consider a local, prevalence-based spread of fear of the disease as a factor that influences the behavior change [22]. We add an unreported compartment to account for individuals who were undetected due to inadequate testing and tracing, or unwillingness to be detected. The addition of an unreported compartment has been used in estimating unreported COVID-19 cases in various countries [23, 24, 25, 26]. It is worth noting these models did not consider behavior change similar to our approach. This study aims to investigate how the behavior and attitude of the people towards COVID-19 during the early phase of the pandemic impact the spread of the disease. In particular, we are interested on the effects of reporting and behavior on the timing and magnitude of the peak of COVID-19 infections in Metro Manila, Philippines from March to September 2020. Furthermore, we present an optimization approach that allows easing from ECQ to GCQ at an earlier time and minimizes the number of additional beds necessary to ensure sufficient capacity in healthcare facilities. Since infectious diseases are likely to emerge or reemerge, there is a need to improve existing policies and plan effective strategies to minimize socioeconomic impact caused by strict community quarantine protocols and ensure that healthcare service is available to those who may need it.

## 2. Methods

### 2.1. Data

The number of cumulative confirmed cases from March 8 to September 30, 2020 were obtained from the Philippine Department of Health (DOH) data drop [6]. These data were used to estimate the rates of transmission, behavior change, and reporting. The data on COVID-19 bed capacity and occupancy rate in Metro Manila were gathered from the number of occupied and available isolation, ward, and ICU beds from the weekly DOH bulletin from June 20 to September 26, 2020 [33]. The total population of Metro Manila was set to 13484462, based on the 2020 census data of the Philippine Statistics Authority [8].

### 2.2. Mathematical model

The model we present is an extension of the model in [21] wherein an unreported compartment is added to represent the undetected or unreported COVID-19 cases in the early-phase of the pandemic in the Philippines. We consider seven compartments: susceptible (*S*), behavior-changed susceptible (*S*_*F*_), exposed (*E*), reported infectious (*I*), unreported infectious (*I*_*u*_), isolated (*Q*), and recovered (*R*). A schematic diagram of the model is shown in Figure 1.

**Figure 1:**
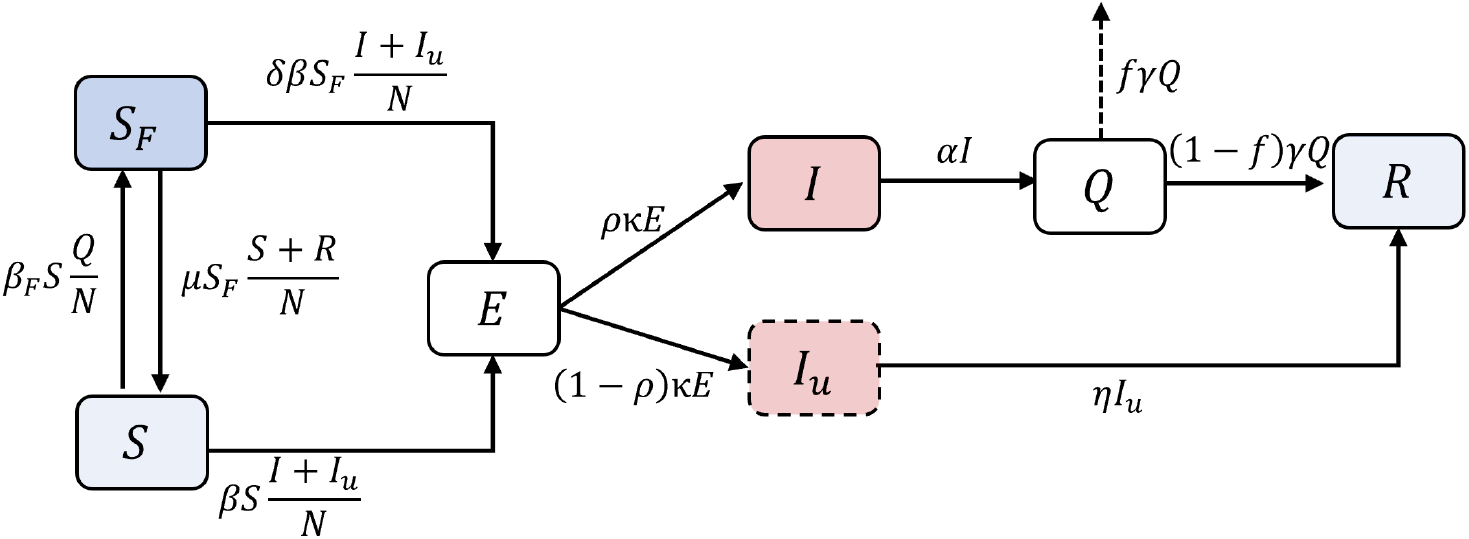
COVID-19 transmission model that incorporates behavior change and unreported cases. Susceptible (*S*) may change behavior (*S*_*F*_) and vice versa at rates *β*_*F*_ or *µ*. These classes can be exposed (*E*) to the virus and become infectious (*I, I*_*u*_) in 1*/κ* days on average. Transmission rate *β* is reduced by a factor *δ* for behavior-changed *S*_*F*_. Reporting ratio is denoted by *ρ*. Confirmed cases are isolated (*Q*) in 1*/α* days and recover (*R*) 1*/γ* days on average. The average fatality rate is denoted by *f*. Those in *I*_*u*_ recover 1*/η* days on average.

Assuming a local, prevalence-based spread of the fear of the disease and following the study of Perra et al. in [22], the transition rate of a susceptible to a behavior-changed susceptible is given by *β*_*F*_ *Q*(*t*)*/N* (*t*). This means that a susceptible is more likely to change behavior as the number of confirmed cases among one’s contacts increase. Moreover, the movement back to *S* is assumed to be influenced by the number of recoveries and susceptible individuals without behavior change [22]. As the recoveries and susceptible individuals increase among the contacts of a behavior-changed susceptible, the more likely the individual to exit the *S*_*F*_ class and resume regular social behavior. The parameter *µ* represents the rate of the easing of behavior and its value is assumed to be 1/14 [21].

Susceptible individuals (*S* and *S*_*F*_) move to the exposed class upon contact with infectious individuals (*I* and *I*_*u*_) at a rate *β*. The transmission rate for the behavior-changed susceptible class is assumed to be reduced by a factor *δ*. The reporting ratio *ρ* partitions the exposed class to reported *I* and unreported *I*_*u*_ classes. Assuming that an individual becomes infectious 2 days before symptom onset [27], mean incubation period of the original virus strain is 6 days [28], and mean duration between symptom onset and the first medical consultation in the Philippines during this time was 6.75 days [29], we set the mean latent period (1*/κ*) to 4 days and mean infectious period of reported cases (1*/α*) to 8.75 days. From isolation, individuals recover 1*/γ* days on average or die. The average fatality rate, which is the ratio of the daily deaths to daily active cases and denoted by *f*, is set to 1.9% [30]. Those in the unreported class are assumed to have less severe symptoms and move to the recovered class 1*/η* days on average. The model equations and the table of parameters are in Appendix A.

### 2.3. Least-squares fitting of parameters

The values of the transmission rates, reporting ratio, and reduction factor were estimated from the cumulative cases data in Metro Manila from March 8 to September 30, 2020. We divide the period into two: period 1 is from March 8 to May 31, while period 2 is from June 1 to September 30, 2020. Metro Manila was mostly under ECQ during period 1, while it was mostly under GCQ during period 2. It was during period 2 that the first major epidemic wave in the Philippines occurred. Since the intensity of NPIs and behavior of the population during ECQ and GCQ vary, the values for the transmission rates (*β* and *β*_*F*_) and reporting ratio (*ρ*) in periods 1 and 2 are assumed to be different. The reduction in transmission (*δ*) for the behavior-changed susceptible class is assumed to have the same value in the two periods. We denote the transmission rates and reporting ratio for periods 1 or 2 by the subscripts 1 or 2, respectively.

Estimation of the parameters was done by fitting the model to the cumulative confirmed cases data at corresponding time points using a least squares approach. That is, we minimize

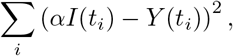

where *Y* (*t*_*i*_) is the total reported cases on day *t*_*i*_. We utilize the Matlab built-in function lsqcurvefit to obtain the parameter estimates for the best model fit.

### 2.4. LHS-PRCC and parameter bootstrapping

Sensitivity analysis is a numerical technique that is widely used in identifying and ranking critical parameters to a model output [31]. A parameter is said to be influential to an output if small perturbations of its value lead to significant changes in the output of the model. In this work, we use Partial Rank Correlation Coefficient (PRCC) method paired with the Latin Hypercube Sampling (LHS) technique. LHS-PRCC is one of the most efficient global sensitivity analysis techniques. To consider every infection, we use the cumulative number of infected individuals *κE* as the model output. In the implementation of LHS, we sampled 10000 combinations of the parameters, all following a uniform distribution. PRCC values of each parameter are calculated at five time points: April 19, May 31, August 2, October 4, and November 1, 2020.

Parameter bootstrapping is a statistical technique to quantify uncertainty and construct confidence intervals of estimated parameters. In this study, we utilize the algorithm introduced in [32], where large samples of synthetic data sets using the estimated model parameters are generated assuming a certain probability distribution structure. In our simulations, parameters are re-estimated from 10000 synthetic data sets each generated by assuming a Poisson error structure. The mean, standard deviation, and 95% confidence intervals of the re-estimated parameters are determined.

### 2.5. Optimization problem

Using the bed occupancy data from the DOH [33], we calculated that an average of 16% of the active cases *Q*(*t*) occupied COVID-19 beds from June to September 2020. Fitting the weekly data on available beds, a linear function representing 75% of the bed capacity was obtained. The DOH categorizes a facility as high risk if the bed occupancy is 70% to 85%, and critical if bed occupancy is greater than 85%. From July 18 to August 8, 2020, most facilities in Metro Manila was on critical or high-risk, with a combined COVID-19 bed occupancy (isolation, ward, and ICU beds) exceeding 75% of the capacity. Here, we propose an optimization approach to determine the number of additional beds per day so that the number of cases requiring beds does not exceed 75% of the total bed capacity and if it is possible to transition from ECQ to GCQ earlier than June 1, 2020.

We denote by *Q*_*H*_ (*t*) the number of active cases requiring beds and *H*(*t*) the linear, time-dependent, data-fitted bed capacity. We consider two objectives:

i. minimize the number of additional hospital beds needed per day (*ω*) so that the 75% capacity is not reached, and
ii. determine an earlier timing (*τ*) of easing from ECQ to GCQ.

The problem can be formulated as a bi-objective constrained optimization problem expressed as follows:

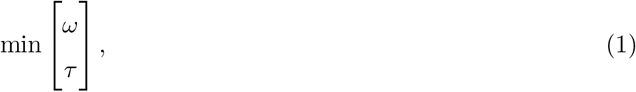

such that

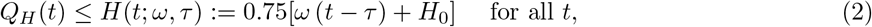

where *Q*_*H*_ (*t*) = 0.16 *· Q*(*t*), *H*_0_ is the baseline number of beds at time *τ* based on the data, and *Q*(*t*) is solved from (A.1) by setting *τ* as the day when GCQ started. Since this is a bi-objective optimization problem, the solution is not unique but a pareto optimal set. We solve (1)-(2) using Genetic Algorithm, which has found a growing number of applications in various fields of science and engineering [34, 35, 36]. In particular, we implement the Matlab built-in function gamultiobj, which is based on a variant of Non-dominated Sorting Genetic Algorithm II (NSGA-II) [37, 38].

## 3. Results

### 3.1. Parameter Estimation

The best model fit for the cumulative and daily cases are shown as the black curves in Figure 2. The red circles represent the data points. The estimated transmission rates *β*_1_ and *β*_*F*,1_ in Period 1 were 0.199 and 471.057, respectively. In Period 2, the transmission rate of the disease *β*_2_ increased to 0.361, while the transmission rate of awareness or fear of the disease *β*_*F*,2_ dropped to 67.783. The reporting ratio *ρ*_1_ in Period 1 was estimated at 28%, which increased to 86% in Period 2. The reduction in transmission induced by behavior change *δ* was estimated at 0.202. The parameter estimates are given in Table A.1.

**Figure 2:**
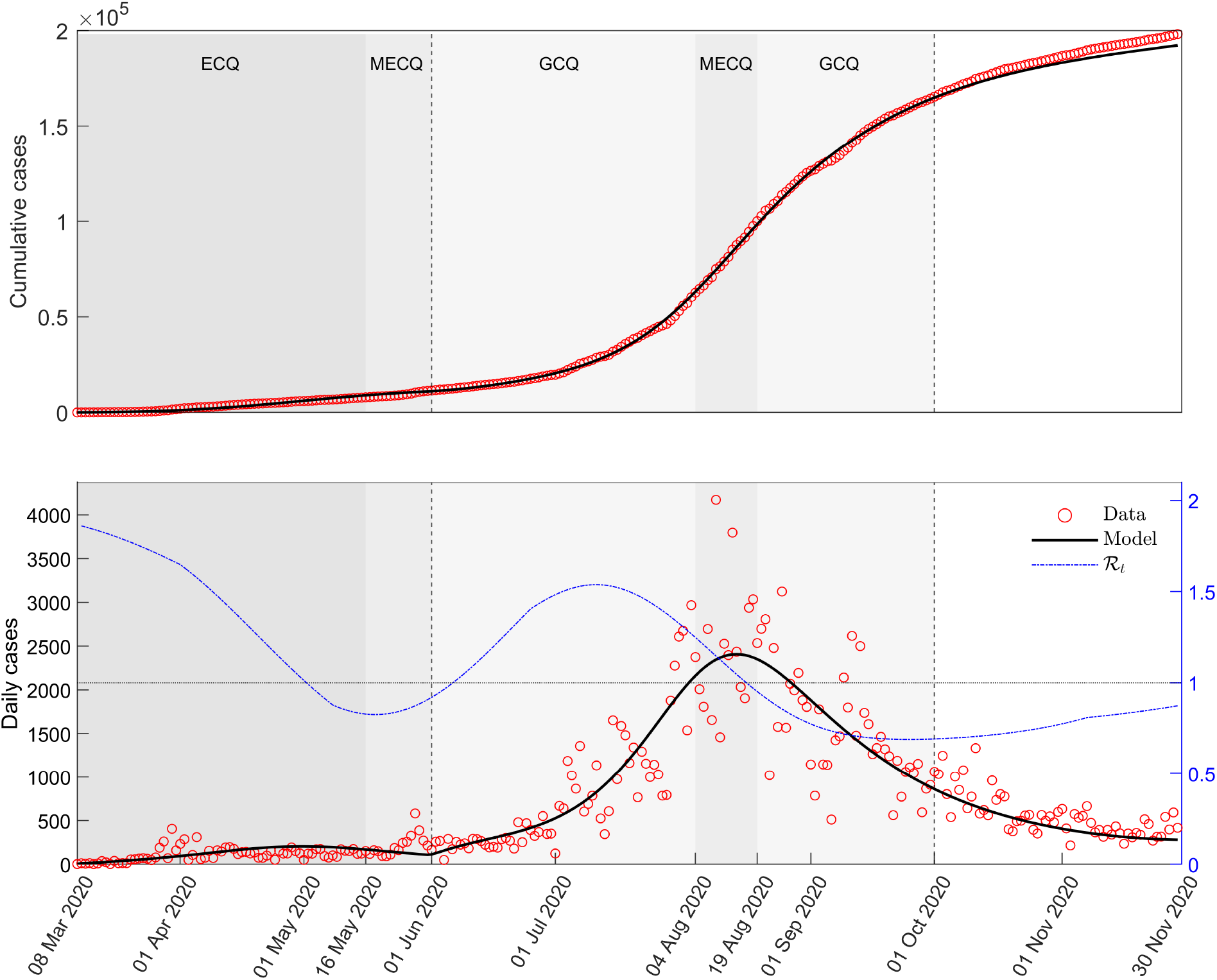
Best model fit to the cumulative cases from March 8 to September 30, 2020. The black curves are the plots of the cumulative cases (top) and daily cases (bottom) using the model and parameter estimates. The vertical dashed lines mark the end of Periods 1 and 2. Metro Manila was under ECQ from March 8 to May 15, 2020 (dark gray), MECQ from May 16 to May 31 and August 4 to 18, 2020 (gray), and GCQ from June 1 to August 3 and August 19 to September 30, 2020 (light gray). The red circles represent the data points and the blue curve is the reproductive number ℛ(*t*).

The reproductive number ℛ(*t*), which is the average number of secondary infections from an individual during one’s infectious period, is shown as the blue curve in Figure 2. Using the next-generation matrix approach [39], it is expressed as

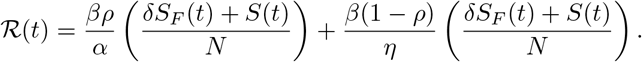

Initially, *ℛ*(*t*) was at 1.9 then decreased to 0.9 by the end of Period 1. During Period 2, *ℛ*(*t*) remained above 1 from early June until mid-August, with peak of up to 1.5 in mid-July 2020.

Using the cumulative number of infected individuals *κE* as the model output, results of the sensitivity analysis showed that *β* (range: 0.82 to 0.92) and *δ* (range: 0.52 to 0.68) have the highest PRCC values, followed by *ρ* (range: −0.49 to −0.44) and the parameters for the infectious periods, *α* (range: −0.48 to −0.46) and *η* (range: −0.45 to −0.28). PRCC values of *κ* declined over time, with its highest value at 0.436. The rest of the parameters have small magnitudes of PRCC. Moreover, the parameter bootstrapping results showed that the re-estimated values of *β, β*_*F*_, *δ* and *ρ* follow a normal distribution and the mean values of the estimates all fall within their respective 95% confidence intervals. Bar plots of the PRCC at different time points are shown in Figure B.6 and the distributions of the re-estimates are shown in Figure B.7 in the Appendix.

### 3.2. Effects of Behavior Change and Reporting

The solid curves in the upper panel of Figure 3 are the plots of the susceptible class *S*, while the dashed lines are the behavior-changed susceptible class *S*_*F*_. Here we investigate what happens if people changed their behavior one (orange), two (yellow), three (purple), or four (green) weeks earlier. We calculated that at the beginning of Period 2, the proportion of *S*_*F*_ with respect to the total susceptible population was 88% (*S*_*F*_ : 11849000; *S*: 1592900). To incorporate early behavior change, we scale the value of *β*_*F*_ to yield the same proportion of *S*_*F*_ one to four weeks before the start of GCQ. The black curves in Figure 3 represent the plots of the model using the parameters in Table A.1.

**Figure 3:**
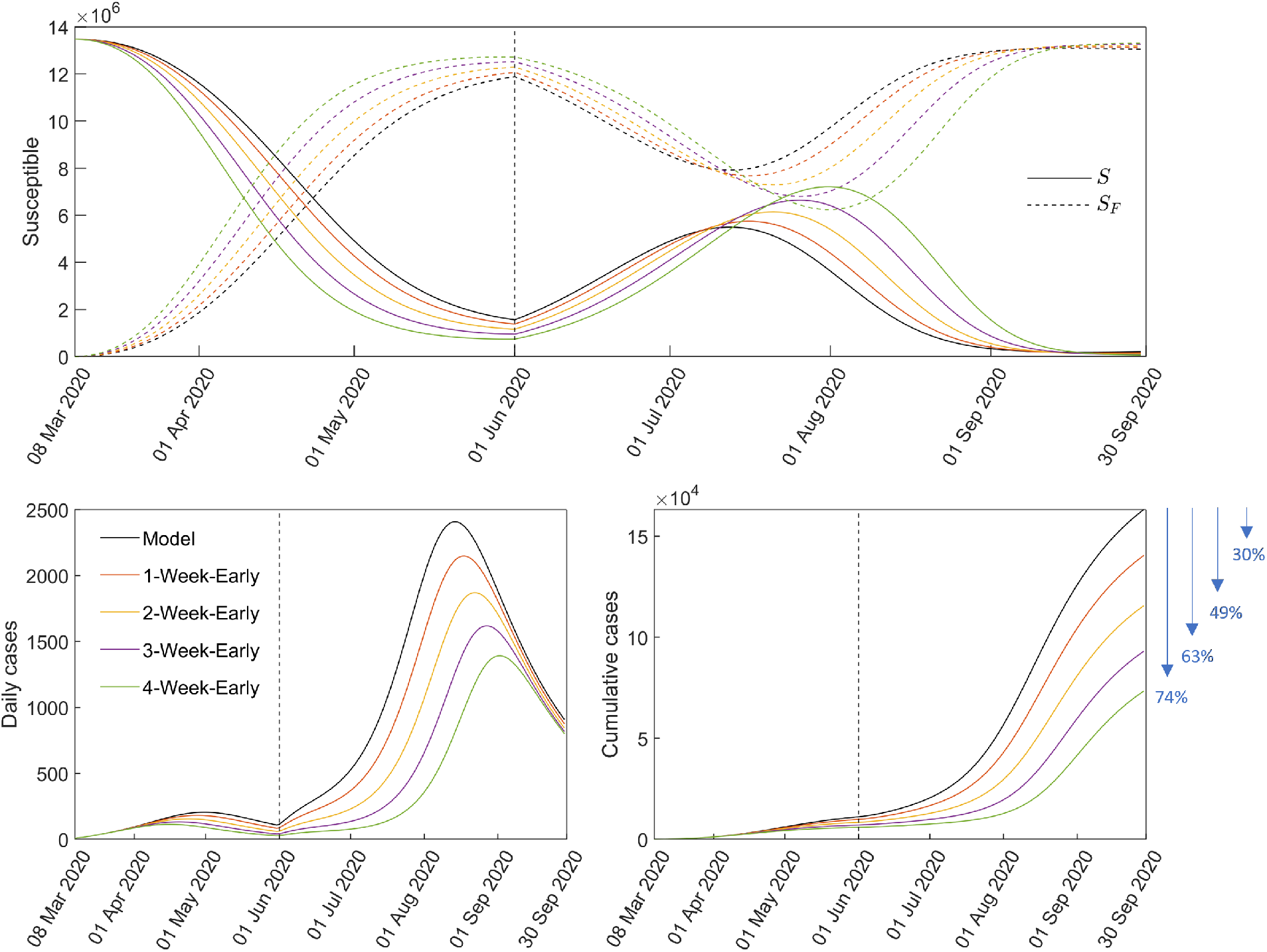
Dynamics of the susceptible population (top panel; *S* solid and *S*_*F*_ dashed curves), daily (lower left panel), and cumulative cases (lower right panel) if the population changed their behavior one (orange), two (yellow), three (purple), or four (green) weeks earlier than the start of Period 2.

During Period 1, we observe a switch in the populations of *S*_*F*_ and *S*. By the end of Period 1, *S*_*F*_ comprises the majority (88%) of the susceptible classes. The impact of early behavior change is seen in the daily and cumulative cases in Period 2, shown in the bottom panels of Figure 3. As behavior changed one, two, three, or four weeks earlier, the cumulative cases decreased to 140468, 115573, 93041, or 73328 from 163191 (model, black), respectively. These translate to reductions of 30%, 49%, 63%, or 74% in the cumulative cases by September 30, 2020. The peak of the daily cases also reduced to 2148, 1870, 1618, and 1392 from 2408 (model, black) and the timing was delayed from one to four weeks.

When the community quarantine was relaxed to GCQ, we observe that the size of *S*_*F*_ declined, while *S* increased. Around mid-July, when the number of daily cases were increasing (*ℛ*(*t*) *>* 1), the behavior of the susceptible classes switched again. From that point until the peak of the first big wave in Metro Manila (∼August 14 according to the model), the proportion of *S*_*F*_ among the susceptible classes increased from 59% to 89%.

The upper panels in Figure 4 show the effect of varying the values of *µ* and *β*_*F*_ on the cumulative cases and timing of the peak of infections. Higher values of *β*_*F*_ and lower factors of the behavior change ease rate *µ* in Period 2 result to notable reductions in the number of cases and delay in the occurrence of the peak. For instance, if in Period 2 we set *β*_*F*,2_ as 3 times *β*_*F*,1_ and *µ* reduced by 90%, then the cumulative cases by the end of September 2020 would have been approximately 30000 and the peak would have occurred around July 25 (140 days from March 8). The blue area on the heatmap for peak timing indicates that the big wave in Period 2 did not occur until September 2020.

**Figure 4:**
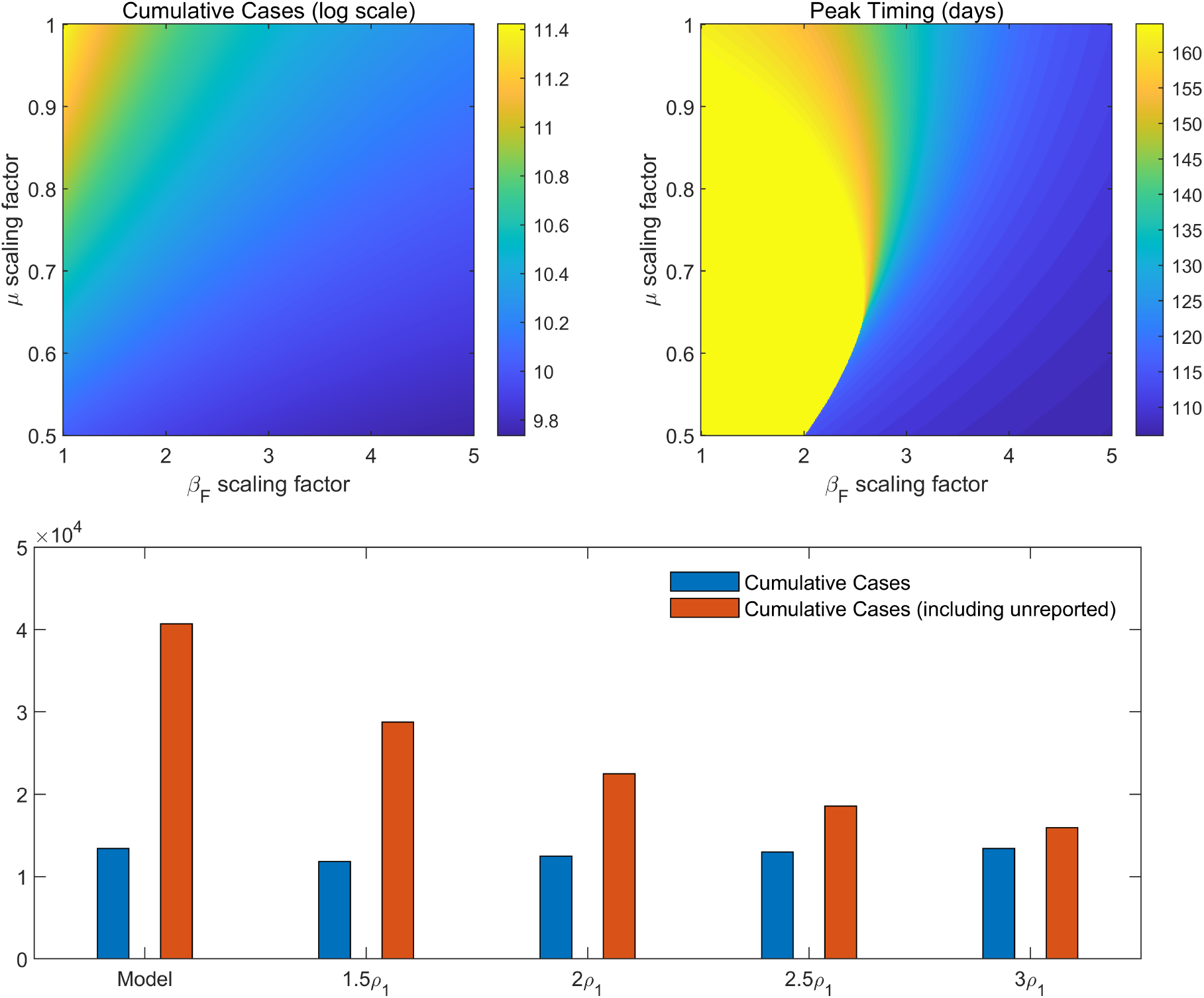
Effect of varying the behavior parameters (*µ* and *β*_*F*_) in Period 2 and reporting ratio in Period 1 (*ρ*_1_) to the timing of the peak and cumulative cases by September 30, 2020.

Finally, the bottom panel in Figure 4 shows the effect of increasing the reporting ratio *ρ*_1_ in Period 1 on the cumulative cases by September 30, 2020. Only slight differences in the reported cumulative cases (blue bars; range: 11817 to 13421) were observed if *ρ*_1_ was increased by factors of 1.5, 2, 2.5, or 3. On the other hand, cumulative cases, including the unreported (red bars), can be reduced by 29%, 45%, 54%, or 61%, if *ρ*_1_ was increased by factors of 1.5, 2, 2.5, or 3, respectively.

### 3.3. Optimal bed capacity and timing of policy change

Panel (A) in Figure 5 shows the fitted bed capacity *H*(*t*) (dashed curve) from the data (red circles) and the number of cases requiring beds *Q*_*H*_ (*t*) from the model. Note that the red circles depict 75% of the total COVID-19 bed capacity during this time and *Q*_*H*_ (*t*) is 16% of *Q*(*t*), representing the average number of active cases that occupy beds. By calculating the slope of *H*(*t*), denoted by *ω*_data_, an estimated 33 beds were added per day from June 21 to September 30, 2022 in Metro Manila. Notably, *Q*_*H*_ (*t*) *> H*(*t*) during the second MECQ, when healthcare workers demanded a ‘timeout’. The peak of *Q*_*H*_ (*t*) was calculated at 5307 cases.

**Figure 5:**
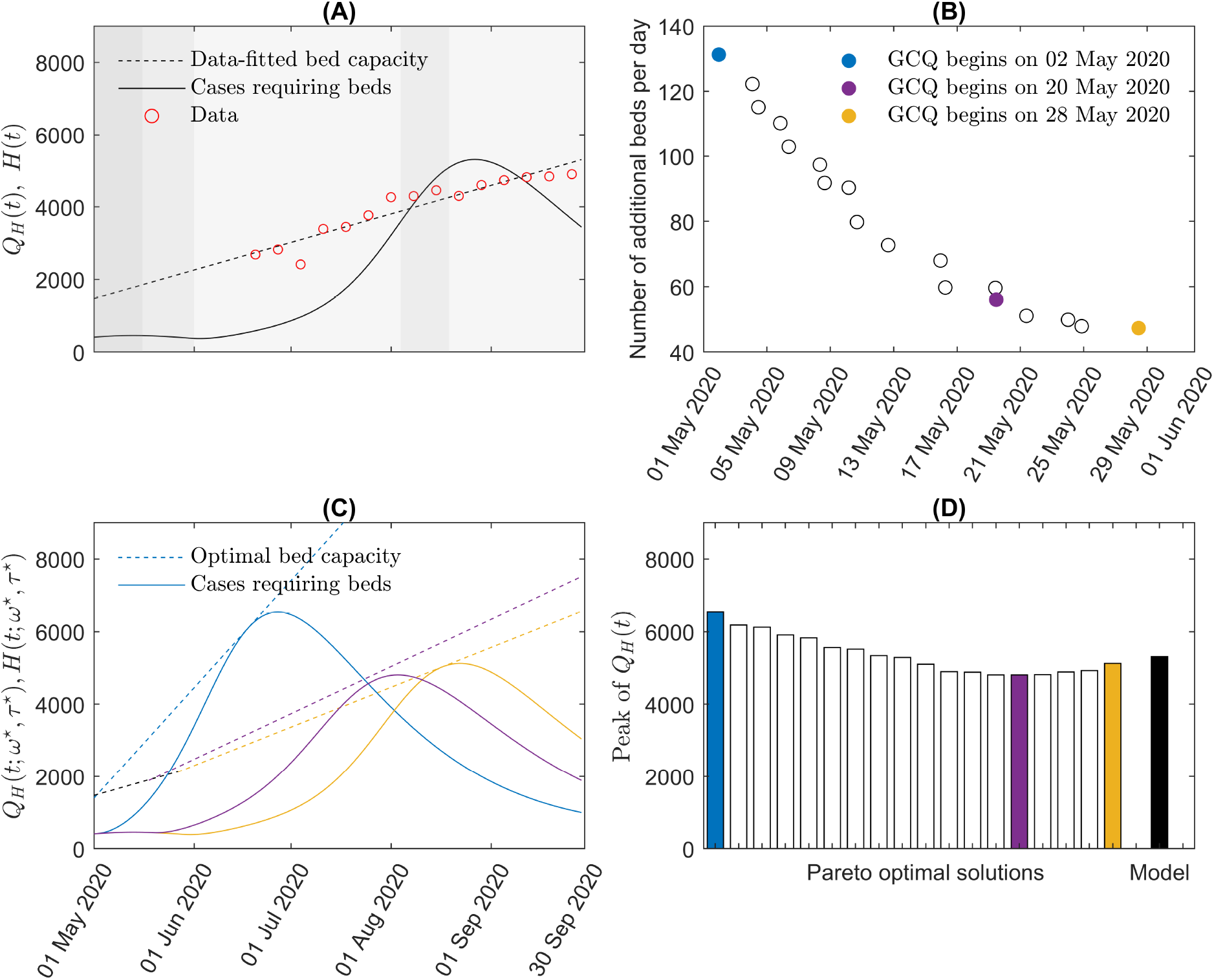
Pareto optimal solutions of the bi-objective optimization problem. (A) The black curve *Q*_*H*_ (*t*) is the number of cases requiring beds, calculated as 16% of the reported active cases *Q*(*t*) and the black dashed line *H*(*t*) is the bed capacity obtained by fitting the data (red circles) using linear regression. (B) Pareto optimal set of (1). (C) Plots of *Q*_*H*_ (*tω*^⋆^, *τ* ^⋆^) (cases requiring beds) and the optimal hospital bed capacity *H*(*t*; *ω*^⋆^, *τ* ^⋆^) corresponding to the three Pareto optimal solutions colored blue, purple, and yellow. (D) Peaks of *Q*_*H*_ (*t*; *ω*^⋆^, *τ* ^⋆^) corresponding to the Pareto optimal solutions, compared to the model.

The optimal solutions of the bi-objective optimization problem in (1) form a Pareto optimal set illustrated in Figure 5 Panel (B). The circles are all optimal solutions depicting different policies. The blue-colored optimal solution corresponds to the earliest easing to GCQ on May 2, 2020 and requires 131 additional beds per day to ensure that the bed capacity is adequate (up to 75% occupancy) during the surge in cases following the lifting of restrictions (blue curves in Panel (C)). On the other hand, the yellow-colored optimal solution has the least number of additional beds per day (47 beds) and latest start of GCQ (on May 28, 2020). This solution has a delayed and lower peak of infections compared to the blue Pareto optimal solution (see Panel (C)). Compared to the curves in Panel (A), the number of cases requiring beds *Q*_*H*_ (*t*; *ω*^⋆^, *τ* ^⋆^) shown in Panel (C) is below the optimal bed capacity *H*(*t*; *ω*^⋆^, *τ* ^⋆^). Hence, constraint (2) of the optimization problem is satisfied. Panel (D) shows the peak sizes of the epidemic waves corresponding to the various Pareto optimal solutions. The smallest peak size (4807 cases, purple) is the optimal solution with GCQ starting on May 20, 2020 and with at least 56 additional beds.

## 4. Discussion

Using the estimated parameter values, we observe that the model captures the trend of the daily and cumulative data from March until November 2020. The model shows a small peak in the number of daily cases (204 cases) and a slow increase in cumulative cases during Period 1. A much higher peak (2408 cases) around mid-August 2020 and a sharp rise in cumulative cases is seen from the model during Period 2. A delay of about one month between the drop in *ℛ*_*t*_ and decline in daily cases was also observed. Results of the parameter bootstrapping suggest good reliability of the estimated parameters. Moreover, sensitivity analysis showed that the transmission rate *β* was the most sensitive parameter with respect to the number of cumulative infections. Higher reporting ratio *ρ* or shorter mean infectious period of reported cases 1*/α* reduces the cumulative infections. These results suggest that intensifying testing and tracing efforts can effectively reduce new infections. The average latent period (1*/κ*), which has the effect of delaying infection, becomes less sensitive to the cumulative number of infections as the epidemic progresses, while the mean infectious period of unreported cases (1*/η*) becomes more sensitive as the epidemic progressed.

Reporting was low in the early pandemic phase, possibly resulting from low testing capacity, slow contact tracing, uncertainty and lack of knowledge about the disease and protocols, or fear of social stigma. This changed during Period 2, where the estimated reporting ratio went up three times. These results are consistent with a study on time-varying under-reporting estimates in various countries, including the Philippines, during the same period [40]. The impact of the community quarantines imposed by the government are reflected on the reduction parameter *δ* and transmission rates *β* and *β*_*F*_. The estimated 20% reduction in transmission for the behavior-changed class compares with a mathematical model of COVID-19 transmission in the Philippines which showed that the *minimum health standards* reduced the probability of transmission per contact by 13 − 27% [41]. As the community quarantine was relaxed from Period 1 to 2, the transmission rate of the disease (*β*) increased from Period 1 to 2, while the rate of behavior change or transmission rate of awareness or fear of COVID-19 (*β*_*F*_) decreased from Period 1 to 2.

Results in Figures 3 and 4 emphasize the importance of early public health campaigns and positive behavior changes (e.g mask wearing, improved hygiene practices, and social distancing) on reducing and delaying the peak of infections. Although fear and stigma can influence behavior changes [42, 43], these can also affect reporting and negatively impact disease control. We see in Figure 4 that as reporting is increased, total cumulative cases including the unreported decreased significantly.

Without vaccines and antiviral therapy, control of epidemic diseases rely on effective NPIs and management of healthcare systems. The bi-objective optimization approach can be used as a decision support tool because of the multiple optimal solutions provided by the method, wherein policymakers can choose depending on how much priority given on minimizing the number of additional beds or earlier easing of restrictions. Although the method is applied to the Philippines, the optimization approach can also be used by other cities or countries by adapting location-specific epidemiological parameters. For countries with limited resources, the solutions corresponding to later easing of restrictions and less number of additional beds may be better options. On the other hand, for those that can provide sufficient additional beds, the approach can be used to identify optimal timing of adjusting NPIs. Results in Figure 5 suggest that if Metro Manila eased to GCQ on June 1, 2020, at least 47 beds per day should have been prepared so that the bed occupancy in the capital did not reach critical or high-risk, and MECQ was not needed to be reimposed. The blue solutions in Figure 5 prioritizes the earlier timing (*τ*) of easing of protocols over the number of additional beds (*ω*). With this policy, GCQ could have been started 30 days earlier. However, this requires 131 additional beds per day, which is 4 times *ω*_data_. On the other hand, the yellow solutions correspond to implementing GCQ on May 29, 2020. This would require 47 additional beds per day, which is still more than *ω*_data_. In Panel (D), the policy in purple has the lowest peak among all the Pareto solutions. For this policy, even though GCQ starts on May 20 (12 days earlier than what happened), the peak of cases (purple curve in Panel (C)) was 500 less than the peak from the model (black curve in (A)). This policy would have required 56 beds per day, which is almost double than *ω*_data_.

## 5. Conclusion

In this work, we used an SEIQR model that considers behavior change and underreporting to study the spread of COVID-19 during the early phase of the pandemic in Metro Manila, Philippines. Behavior change can be influenced by awareness or fear of the disease, and willingness to observe NPIs such as social distancing and mask-wearing. It was incorporated to the model by introducing a two-compartment susceptible population: one for the behavior-changed population and the other for those with regular behavior. The probability of getting infected is reduced for the behavior-changed susceptible class. Due to limited testing and tracing, or negative attitudes of people towards seeking healthcare, a compartment for the unreported cases was also added.

The results of this study highlight the importance of early behavior change and high reporting rate in reducing the number of cases and delaying the peak of infections. These can be done by intensifying case surveillance and public health campaigns promoting compliance to NPIs, seeking healthcare, and discouraging social stigma. Moreover, this study provides an optimization approach that quantifies the additional bed requirement when policies are eased. The approach can be helpful in planning strategies that address strengthening or easing of policies, especially during the early phase when NPIs were the only control measures. Although the study focused on the transmission of COVID-19 in the Philippines, the proposed model is general enough that it can be applied to any city or country. The optimization problem can also be applied to other disease outbreaks by adjusting key epidemiological parameters. A limitation of the model is that it can only describe the early phase of the epidemic, and so for future work, the model can be extended to describe succeeding epidemic waves and incorporating vaccination, variants, attitudes and behavior of the people towards the disease, and actions done by government. Since the simulations did not consider the economic impact of NPIs, a model that maximizes economic output of a city or country while minimizing the number of infections is another work that can be pursued.

## Data Availability

The datasets supporting the conclusions of this article are available in the Department of Health COVID-19 tracker repository in https://doh.gov.ph/covid19tracker and Philippine Statistics Authority Highlights of the National Capital Region (NCR) Population 2020 Census of Population and Housing (2020 CPH) in https://psa.gov.ph/population-and-housing/node/165009.

https://psa.gov.ph/population-and-housing/node/165009

## 6. Declarations

## Acknowledgments

The authors acknowledge Dr. Peter Julian Cayton of the UP Resilience Institute for his assistance with the data collection.

## Author’s contributions

VMPM and RM, Conceptualization, Draft preparation, Methodology, and Software. YK abd JL, Conceptualization, Draft preparation, Methodology. EJ, Conceptualization, Draft preparation, Methodology, Project Supervision. All authors read and approved the final version of the manuscript.

## Disclaimer

The funders played no role in the design, data collection, analysis, interpretation or writing of this study.

## Patient and public involvement

Patients and/or the public were not involved in the design, or conduct, or reporting, or dissemination plans of this research.

## Patient consent for publication

Not applicable.

## Ethics approval

Not applicable.

## Provenance and peer review

Not commissioned; externally peer reviewed.

## Appendix A. Model Equations

The following system of differential equations describe the model used in the study.

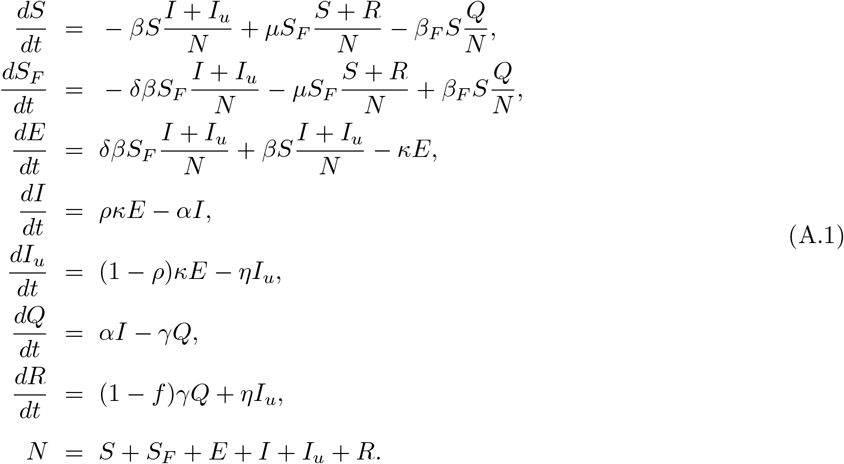

We set the initial number of infectious individuals *I*_0_, exposed *E*_0_, and unreported *I*_*u*,0_ equal to the number of cases 1*/α*, 1*/α* + 1*/κ*, and 10 *× I*_0_ days from March 8, 2020, respectively. The initial number of isolated individuals *Q*_0_ was the number of cases at the start of the estimation period. The initial population of the *S* class is computed by getting the difference between the total population and *E*_0_, *I*_*u*,0_, *I*_0_, and *Q*_0_. The rest of the state variables were initially set to zero. The model parameters and initial values of the state variables are shown in Table A.1.

## Appendix B. Results on sensitivity analysis and bootstrapping

Figure B.6 shows the PRCC for the model parameters with total infections *κE* as the model output. Figure B.7 shows the distributions of the parameter re-estimates. The mean, standard deviation, and confidence intervals are also indicated in the figure.

**Table A.1:** List of model parameters, their values, and references. The subscripts 1 and 2 denote the parameter in Periods 1 and 2, respectively.

| **Symbol** | **Description** (unit) | **Value** | **Ref** |
| --- | --- | --- | --- |
| $\beta_1, \beta_2$ | Transmission rate of COVID-19 in Periods 1 and 2<br>(1/day) | 0.199, 0.361 | Estimated |
| $\beta_{F,1}, \beta_{F,2}$ | Transmission rate of awareness or fear of the disease<br>in Periods 1 and 2 (1/day) | 471.057, 68.783 | Estimated |
| $\mu$ | Behavior change ease rate (1/day) | 1/14 | Assumed |
| $\delta$ | Transmission reduction factor for behavior-changed<br>individuals | 0.202 | Estimated |
| $1/\kappa$ | Mean latent period (day) | 4 | [27, 28] |
| $1/\alpha$ | Mean infectious period of reported cases (day) | 8.75 | [27, 29] |
| $1/\gamma$ | Mean recovery period (day) | 16 | [29] |
| $f$ | Mean fatality rate | 1.9% | [30] |
| $\rho_1, \rho_2$ | Reporting ratio in Periods 1 and 2 | 0.289, 0.866 | Estimated |
| $1/\eta$ | Mean infectious period of unreported cases (day) | 10 | [44] |
| $S_0$ | Initial susceptible population | 13483232 | Calculated |
| $S_{F,0}$ | Initial behavior-changed susceptible population | 0 | Assumed |
| $E_0$ | Initial exposed population | 139 | Assumed |
| $I_0$ | Initial reported infectious population | 99 | Calculated |
| $I_{u,0}$ | Initial unreported infectious population | 990 | Assumed |
| $Q_0$ | Initial isolated population | 2 | [6] |
| $R_0$ | Initial recovered population | 0 | Assumed |

**Figure B.6:**
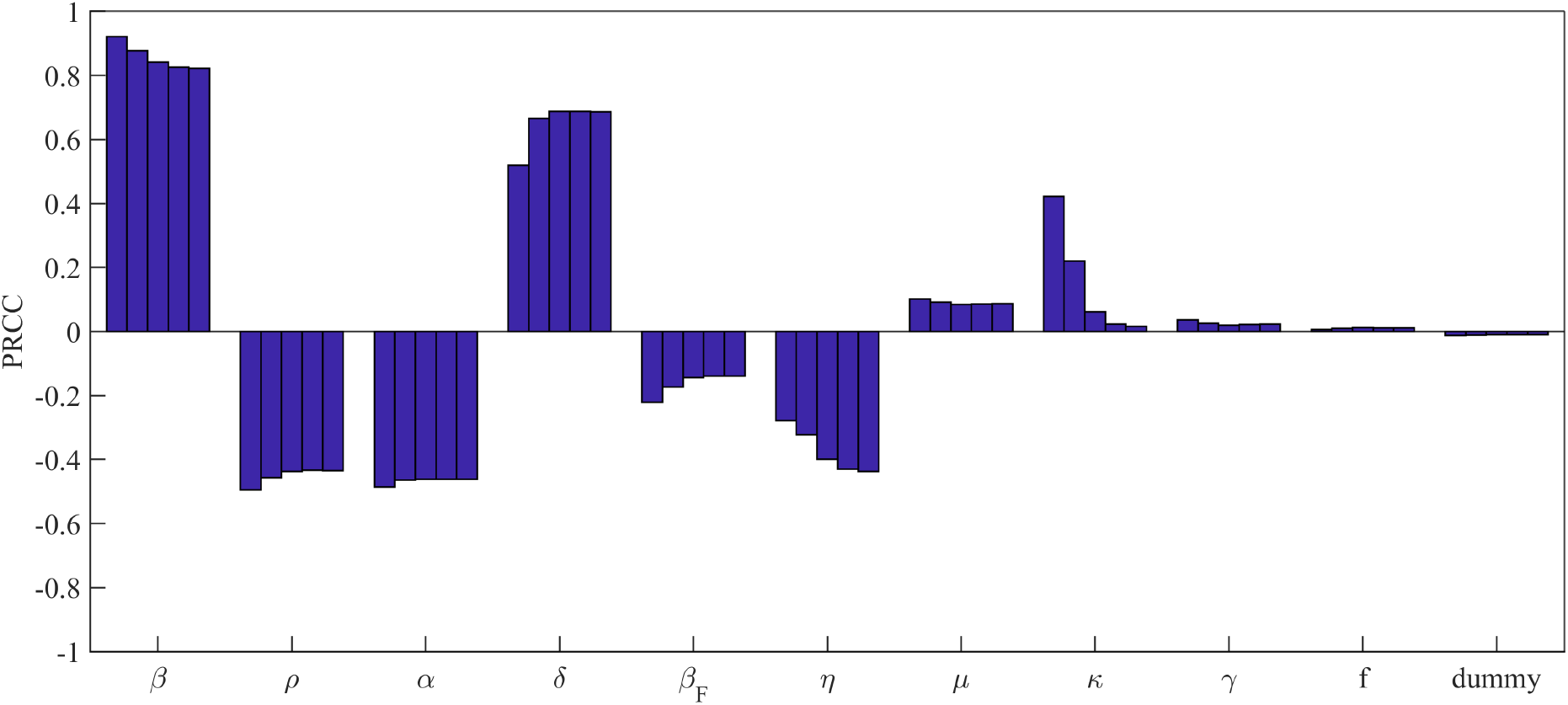
PRCC values of the model parameters, with respect to the cumulative number of infected individuals *κE*. The bars represent the PRCC values on April 19, May 31, August 2, October 4 and November 1, 2020.

**Figure B.7:**
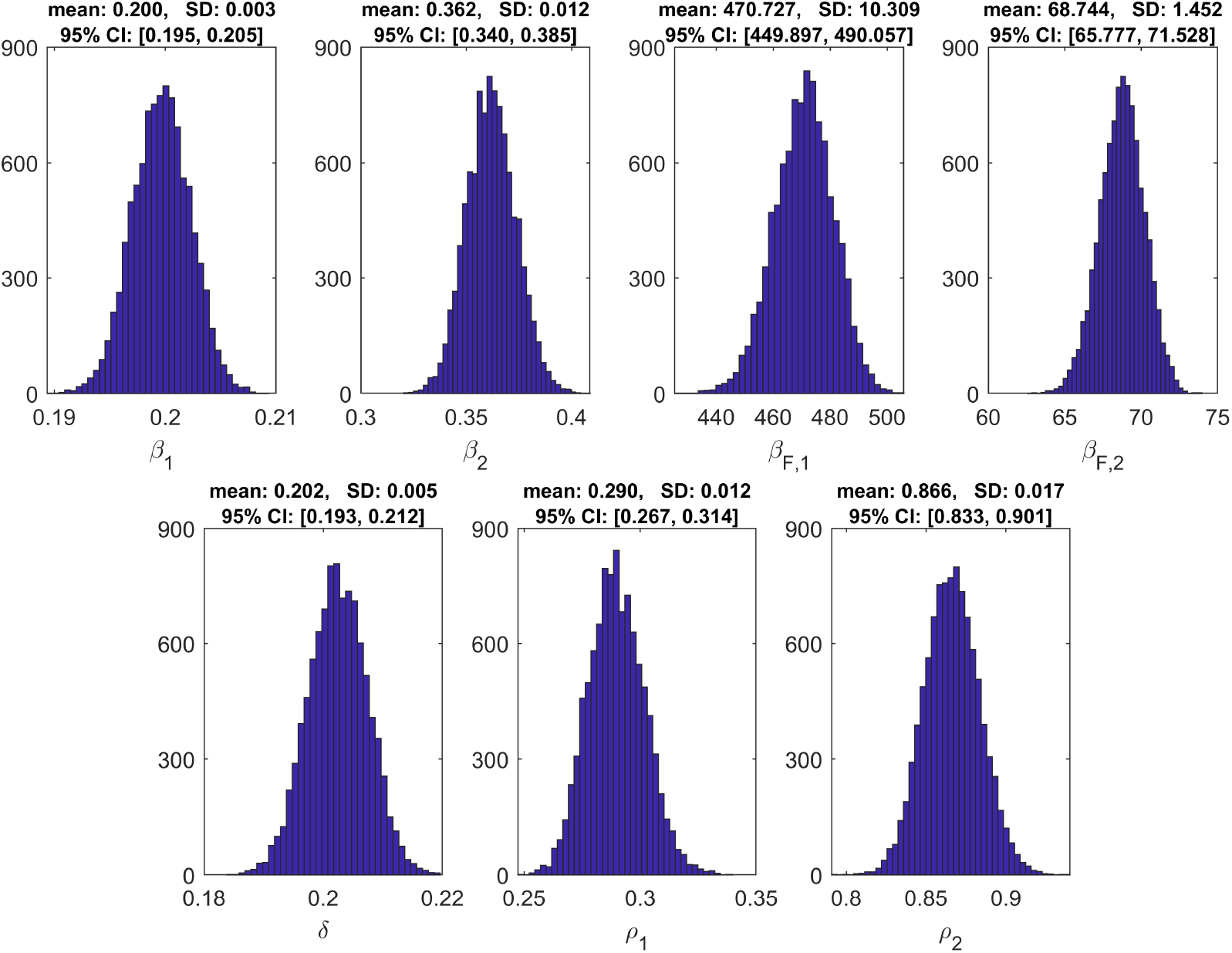
Distribution of the re-estimates of *β*_1_, *β*_2_, *β*_*F*,1_, *β*_*F*,2_, *δ, ρ*_1_ and *ρ*_2_ in the parameter bootstrapping. The mean, standard deviation (SD), and 95% confidence intervals are also shown.

## References

[1] J. Vallejo, Benjamin M., R. A. C. Ong, Policy responses and government science advice for the COVID 19 pandemic in the Philippines: January to April 2020, Progress in Disaster Science 7 (2020) 100115. doi:10.1016/j.pdisas.2020.100115.

[2] Department of Health, COVID-19 Inter-agency Task Force for the Management of Emerging Infectious Diseases Resolutions, Omnibus Guidelines on the Implementation of Community Quarantine in the Philippines (May 15, 2020), accessed: 2022-05-31. URL https://doh.gov.ph/COVID-19/IATF-Resolutions

[3] World Health Organization, COVID-19 in the Philippines Situation Report 12 (March 30, 2020), accessed: 2022-05-31. URL https://www.who.int/philippines/internal-publications-detail/covid-19-in-the-philippines-situation-report-12

[4] B. Magsambol, PH needs 94,000 contact tracers-DOH, accessed: 2022-07-08. URL https://www.rappler.com/nation/philippines-needs-contact-tracers

[5] World Health Organization, COVID-19 in the Philippines Situation Report 55 (September 29, 2020), accessed: 2022-05-31. URL https://www.who.int/philippines/internal-publications-detail/covid-19-in-the-philippines-situation-report-55

[6] Department of Health, COVID-19 Tracker Philippines, accessed: 2022-07-08. URL https://doh.gov.ph/covid19tracker

[7] D. Dowdy, G. D’Souza, COVID-19 Testing: Understanding the “Percent Positive”, accessed: 2022-07-08. URL https://publichealth.jhu.edu/2020/covid-19-testing-understanding-the-percent-positive

[8] P. S. Authority, 2020 Census of Population and Housing (2020 CPH) Population Counts Declared Official by the President, accessed: 2022-06-14. URL https://psa.gov.ph/content/2020-census-population-and-housing-2020-cph-population-counts-decla

[9] Department of Health, COVID-19 Inter-agency Task Force for the Management of Emerging Infectious Diseases Resolution No. 13 (March 17, 2020), accessed: 2022-05-31. URL https://doh.gov.ph/COVID-19/IATF-Resolutions

[10] Department of Health, DOH Case Bulletin No. 149 (August 10, 2020), accessed: 2022-06-14. URL https://doh.gov.ph/node/23979

[11] S. Tomacruz, After frontliners’ plea, Duterte reverts Metro Manila to MECQ starting August 4, accessed: 2022-07-12. URL https://www.rappler.com/nation/after-frontliners-plea-duterte-reverts-metro-manila-mecq-starti

[12] Department of Health, COVID-19 Inter-agency Task Force for the Management of Emerging Infectious Diseases Resolution No. 64 (August 17, 2020), accessed: 2022-06-16. URL https://doh.gov.ph/COVID-19/IATF-Resolutions

[13] L. L. Lau, N. Hung, D. J. Go, M. Choi, W. Dodd, X. Wei, Dramatic increases in knowledge, attitudes and practices of COVID-19 observed among low-income households in the Philippines: A repeated cross-sectional study in 2020, Journal of Global Health 12.

[14] K. Hapal, The Philippines’ COVID-19 response: Securitising the pandemic and disciplining the pasaway, Journal of Current Southeast Asian Affairs 40 (2) (2021) 224–244.

[15] N. Quijano, M. C. Fernandez, A. Pangilinan, Misplaced priorities, unnecessary effects: Collective suffering and survival in pandemic Philippines, Asia-Pasific Journal: Japan Focus 18 (15) (2020) 1–15.

[16] J. C. G. Corpuz, ‘We are not the virus’: stigmatization and discrimination against frontline health workers, Journal of Public Health 43 (2) (2021) e327–e328.

[17] J. G. S. Kahambing, S. R. Edilo, Stigma, exclusion, and mental health during COVID19: 2 cases from the Philippines, Asian Journal of Psychiatry 54 (2020) 102292.

[18] S. Kim, Y. B. Seo, E. Jung, Prediction of COVID-19 transmission dynamics using a mathematical model considering behavior changes in Korea, Epidemiology and health 42.

[19] J. Lee, S.-M. Lee, E. Jung, How Important Is Behavioral Change during the Early Stages of the COVID-19 Pandemic? A Mathematical Modeling Study, International Journal of Environmental Research and Public Health 18 (18) (2021) 9855.

[20] S. Kim, Y.-J. Kim, K. R. Peck, E. Jung, School opening delay effect on transmission dynamics of coronavirus disease 2019 in Korea: based on mathematical modeling and simulation study, Journal of Korean medical science 35 (13).

[21] S. Kim, Y. Ko, Y.-J. Kim, E. Jung, The impact of social distancing and public behavior changes on COVID-19 transmission dynamics in the Republic of Korea, PLOS ONE 15 (9) (2020) e0238684. doi:10.1371/journal.pone.0238684. URL http://dx.doi.org/10.1371/journal.pone.0238684

[22] N. Perra, D. Balcan, B. Gonçalves, A. Vespignani, Towards a characterization of behavior-disease models, PloS one 6 (8) (2011) e23084.

[23] Z. Liu, P. Magal, G. Webb, Predicting the number of reported and unreported cases for the COVID-19 epidemics in China, South Korea, Italy, France, Germany and United Kingdom, Journal of theoretical biology 509 (2021) 110501.

[24] V. Deo, G. Grover, A new extension of state-space SIR model to account for Underreporting–An application to the COVID-19 transmission in California and Florida, Results in Physics 24 (2021) 104182.

[25] M. Melis, R. Littera, Undetected infectives in the Covid-19 pandemic, International Journal of Infectious Diseases 104 (2021) 262–268.

[26] B. Ivorra, M. R. Ferrández, M. Vela-Pérez, A. M. Ramos, Mathematical modeling of the spread of the coronavirus disease 2019 (COVID-19) taking into account the undetected infections. The case of China, Communications in nonlinear science and numerical simulation 88 (2020) 105303.

[27] World Health Organization et al., Transmission of SARS-CoV-2: implications for infection prevention precautions: scientific brief, 09 July 2020, Tech. rep., World Health Organization (2020).

[28] Y. Wang, R. Chen, F. Hu, Y. Lan, Z. Yang, C. Zhan, J. Shi, X. Deng, M. Jiang, S. Zhong, et al., Transmission, viral kinetics and clinical characteristics of the emergent SARS-CoV-2 Delta VOC in Guangzhou, China, EClinicalMedicine 40 (2021) 101129.

[29] N. J. L. Haw, J. Uy, K. T. L. Sy, M. R. M. Abrigo, Epidemiological profile and transmission dynamics of COVID-19 in the Philippines, Epidemiology and Infection 148. doi:10.1017/s0950268820002137.

[30] L. Rampal, B. Liew, M. Choolani, K. Ganasegeran, A. Pramanick, S. Vallibhakara, P. Tejativaddhana, V. Hoe, Battling covid-19 pandemic waves in six south-east asian countries: A real-time consensus review, Med J Malaysia 75 (6) (2020) 613–625.

[31] S. Marino, I. B. Hogue, C. J. Ray, D. E. Kirschner, A methodology for performing global uncertainty and sensitivity analysis in systems biology, Journal of Theoretical Biology 254 (1) (2008) 178–196. doi:10.1016/j.jtbi.2008.04.011.

[32] G. Chowell, Fitting dynamic models to epidemic outbreaks with quantified uncertainty: A primer for parameter uncertainty, identifiability, and forecasts, Infectious Disease Modelling 2 (3) (2017) 379–398. doi:10.1016/j.idm.2017.08.001.

[33] Department of Health, Beat COVID-19 Today: A COVID-19 Philippine Situationer, accessed: 2022-07-20. URL https://drive.google.com/drive/folders/1Wxf8TbpSuWrGBOYitZCyFaG_NmdCooCa?usp=sharing

[34] X.-S. Yang, Nature-inspired optimization algorithms: Challenges and open problems, Journal of Computational Science 46 (2020) 101104.

[35] S. Katoch, S. S. Chauhan, V. Kumar, A review on genetic algorithm: past, present, and future, Multimedia Tools and Applications 80 (5) (2021) 8091–8126.

[36] S. Sharma, V. Kumar, Application of genetic algorithms in healthcare: A review, Next Generation Healthcare Informatics (2022) 75–86.

[37] K. Deb, Multi-objective optimisation using evolutionary algorithms: an introduction, in: Multi-objective evolutionary optimisation for product design and manufacturing, Springer, 2011, pp. 3–34.

[38] K. Deb, A. Pratap, S. Agarwal, T. Meyarivan, A fast and elitist multiobjective genetic algorithm: Nsga-ii, IEEE transactions on evolutionary computation 6 (2) (2002) 182–197.

[39] O. Diekmann, J. Heesterbeek, M. G. Roberts, The construction of next-generation matrices for compartmental epidemic models, Journal of the Royal Society Interface 7 (47) (2010) 873–885.

[40] T. W. Russell, N. Golding, J. Hellewell, S. Abbott, L. Wright, C. A. Pearson, K. van Zandvoort, C. I. Jarvis, H. Gibbs, Y. Liu, et al., Reconstructing the early global dynamics of under-ascertained covid-19 cases and infections, BMC medicine 18 (1) (2020) 1–9.

[41] J. M. Caldwell, E. de Lara-Tuprio, T. R. Teng, M. R. J. E. Estuar, R. F. R. Sarmiento, M. Abayawardana, R. N. F. Leong, R. T. Gray, J. G. Wood, L.-V. Le, et al., Understanding covid-19 dynamics and the effects of interventions in the philippines: A mathematical modelling study, The Lancet Regional Health-Western Pacific 14 (2021) 100211.

[42] L. L. Lau, N. Hung, D. J. Go, J. Ferma, M. Choi, W. Dodd, X. Wei, Knowledge, attitudes and practices of COVID-19 among income-poor households in the Philippines: A cross-sectional study, Journal of global health 10 (1).

[43] J. Choi, K.-H. Kim, The differential consequences of fear, anger, and depression in response to covid-19 in south korea, International Journal of Environmental Research and Public Health 19 (11) (2022) 6723.

[44] Center for Disease Control and Prevention, Ending Isolation and Precautions for People with COVID-19: Interim Guidance, accessed: 2022-07-08. URL https://www.cdc.gov/coronavirus/2019-ncov/hcp/duration-isolation.html

